# Perceived Stress Among Faculty and First Year Students at a College of Pharmacy

**DOI:** 10.1101/2021.10.22.21265395

**Authors:** Melinda Verdone, Milena Murray, Brooke Griffin, Sally Arif, Jennifer Phillips, Kathy Komperda, Susan Winkler, Annette Gilchrist

## Abstract

**Purpose:** Female faculty and students could be affected by stressors disproportionately compared to male counterparts, especially those with children or family obligations. A study was undertaken to determine: 1) stress levels of pharmacy faculty and first-year pharmacy students; 2) whether gender affected faculty and/or student stress levels disproportionally; and 3) how child and family care responsibilities influenced stress levels.

**Methods:** All first-year (P1) students enrolled in a College of Pharmacy were surveyed along with faculty. Stress levels were assessed using the Perceived Stress Scale (PSS10). Additional demographic information, including items related to children and family obligations, was collected.

**Results:** Faculty reported average perceived stress levels (*M*=15.50) while first-year students reported high perceived stress levels (*M*=21.14). Perceived stress levels of female faculty (*M*=16.43) were higher than those of male faculty (*M*=12.00). Perceived stress levels of female students (*M*=22.60) were higher than those of male students (*M*=16.78). Perceived stress levels of female faculty with younger children (*M*=18.85) were higher than those of male faculty with younger children (*M*=9.67). Perceived stress levels of female students with ≥10 hours of family obligations per week (*M*=22.71) were higher than male pharmacy students with ≥10 hours (*M*=12.80).

**Conclusion:** Lower levels of perceived stress for faculty compared to students may be due to the development of coping strategies coinciding with maturity. Results suggest more time spent on family obligations is negatively associated with stress levels for females, but not males. Colleges of pharmacy should invest resources to help reduce stress levels in faculty and student populations, particularly for the female gender.

## Introduction

Pharmacy school can be a stressful experience for both students and faculty, yet there is a lack of published data outlining pharmacy-specific for the rates of depression, anxiety, or other mental health-related illnesses. On average, it has been reported that two-thirds of pharmacy students exhibit unhealthy levels of perceived stress.^1^ Frequently cited sources of stress for pharmacy students include coursework, grades, lack of sleep, financial concerns, and family/relationship issues.^2,3^

Pharmacy faculty regularly work more than 40 hours per week.^4,5^ A common stressor identified among pharmacy faculty is the lack of work-life balance.^6^ For pharmacy faculty, this conflict between the demands of work and personal life can lead to a decrease in overall life satisfaction.^6^ According to El-Ibiary, Yam, and Lee,^5^ 41.3% of pharmacy practice faculty are categorized as emotionally exhausted using the Maslach Burnout Inventory.

The American Association of Colleges of Pharmacy (AACP) Student Affairs Standing Committee addressed pharmacy student wellness and resilience in a report focusing on consequences of burnout, wellness culture, and creating campus cultures to promote wellbeing.^7^ In 2019, the American Pharmacists Association (APhA), AACP, the Accreditation Council for Pharmacy Education (ACPE), the National Association of Boards of Pharmacy (NABP), and the National Alliance of State Pharmacy Associations (NASPA) provided 50 recommendations on how to enhance wellness via sustainable solutions to create improvements in critical areas related to well-being and resilience.^8^

Prior work has shown that, in general, women are affected by stress to a greater extent than men, and women with children are particularly affected. According to Matud,^9^ women, in general, experience more stressful events associated with health and family, while men report more stressful work and finance events. It has also been reported that women with children experience more negative mood states and cognitive failures compared to men.^10^ When women with children take on career responsibilities, stress levels rise even higher. Women with both work and family responsibilities report higher psychological stress than other groups,^10^ and they report that combining work and family is their greatest source of stress.^11^

With the knowledge that female pharmacy faculty and pharmacy students could be affected by stressors disproportionately compared to their male counterparts, especially if they have child or family obligations, a study was undertaken to determine stress levels of pharmacy faculty and pharmacy students and to discern whether child and family care responsibilities influenced stress levels. To our knowledge, no published studies have compared stress levels of faculty and student populations, nor have they assessed if family roles contribute to stress for these populations. The objectives of this study were to 1) determine the stress levels of faculty and first-year (P1) pharmacy students at a single college of pharmacy, 2) compare stress levels of female faculty and P1 pharmacy students to male faculty and pharmacy students, and 3) establish whether family obligations are a contributing factor to stress for either or both pharmacy faculty and pharmacy students.

## Methods

### Study Design, Participants, and Procedures

The study was conducted at a private, graduate-level, professional health sciences university located in the Midwest before the beginning of the COVID-19 pandemic. For this study, 134 first-year (P1) pharmacy students enrolled in the College of Pharmacy 4 year program were surveyed along with all pharmacy faculty employed at the time. This study was considered exempt by the Midwestern University Institutional Review Board.

Quantitative data for (P1) pharmacy students was obtained through an anonymous survey given to all first year students at MWU. The survey was administered in electronic format via Google Forms at the midpoint of the first year during a required course. Survey completion was optional and did not impact students’ course grades. A link to the website where the survey could be found was projected on-screen and shared through the course’s learning management system site. The first page of the electronic survey was a cover letter providing study details and contact information. The link to the survey remained open for submissions through the end of the day. Questions were included to provide information on demographics (age, gender, ethnicity/racial identity, family care (hours per week), if they were from in/out of state, sleep (hours per day), work (hours per week), student loan debt ($), and GPA.

Quantitative data for faculty was obtained through an anonymous self-administered electronic survey via Survey Monkey. The survey remained open for two weeks, with one reminder email sent. Questions were included to provide information on demographics (gender, marital status, if they had children (age <1, 1-12, 12-18, >18 years), and employment-related issues (department, tenure status, if they had a clinical practice site, if they were teaching this quarter).

### Study Measures

Perceived stress in both faculty and students was measured using PSS10 self-administered questionnaires.^12^ The PSS10 is a modified version of the 14 question Perceived Stress Scale (PSS14).^13^ This widely employed psychological instrument provides a global measure of stress related to the degree to which individuals find their lives uncontrollable, unpredictable, and overloaded in the prior month. The questions are general and do not contain content specific to any subpopulation group. Participants rated each item on a 5-point scale based on the frequency with which a particular event was experienced: 0 (never), 1 (almost never), 2 (sometimes), 3 (fairly often), and 4 (often). Four of the 10 items (items 4, 5, 7, and 8) were designed to identify positive events and were reversed-scored. PSS-10 items are introduced with “In the last month, how often have you felt…” followed by items such as “nervous and stressed” and “that you were unable to control the important things in your life.” PSS10 items are provided in **Figure 1**. Responses were summed to create a psychological stress score, with higher total scores indicating greater psychological stress.^13^ Total scores for this measure fall between 0 and 40. Based on the PSS10 scoring guidelines, scores lower than 13 suggest lower levels of stress, scores from 13-19 represent average stress levels, and scores of 20 or higher show high stress levels.^12^

**Figure 1.**
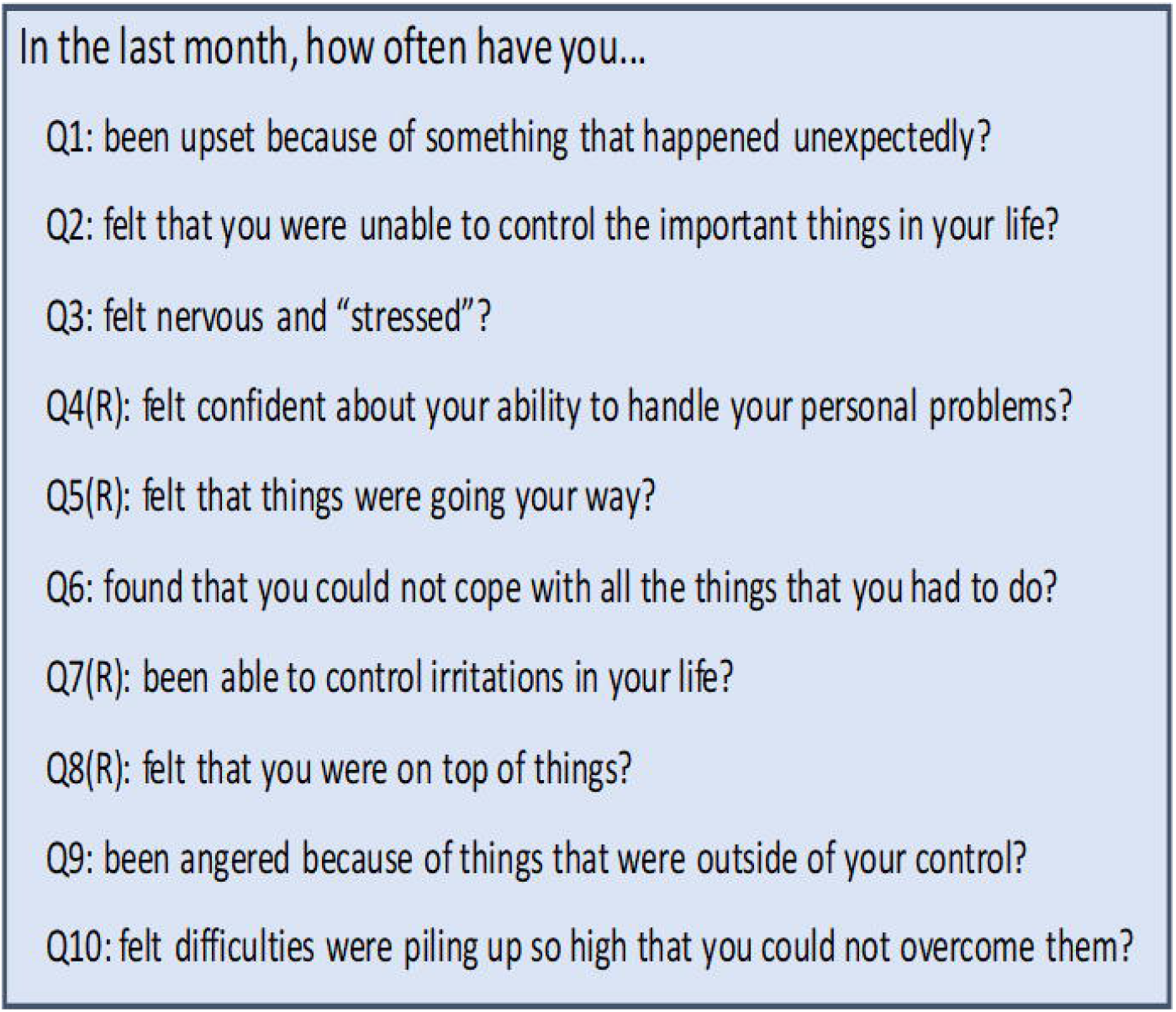
Items on the PSS10 questionnaire provided to first year pharmacy students and pharmacy faculty. The survey started with the sentence “Please rate the following statements related to perceived stress.” Questions 4, 5, 7, and 8 were reverse scored (R).

### Statistical Methods

The collected data underwent quantitative analysis using SPSS (version 26). Descriptive statistics were used to show the data’s central tendency and dispersion. For all analyses, alpha was set *a priori* at 0.05. Independent samples *t*-tests were used to compare PSS-10 scores between faculty and students and between male and female participants within faculty and student groups. A 2 × 2 two-way Analysis of Variance (ANOVA) was performed for both faculty and students to determine the effect of gender and family obligations on perceived stress. For gender, only male and female gender identities were included in the analyses. For family obligations, the demographic question was worded differently between the student and faculty surveys: faculty surveys had a demographic question on whether they had children and, if so, the age (<1, 1 to <12, 12-18, >18 years). Age values were selected based on the premise that infants require more attention than older children, as reflected in child to adult ratios ^14^. Children under 12 years old need more childcare time than those over 12 years of age ^15^ Based upon responses, faculty were ultimately grouped into the following categories: no children, children ≥12 years of age, or children <12 years of age. Students were categorized into two groups: those with ≥10 hours of family care per week and those with <10 hours of family care per week. For students, it has been shown that as few as 10 hours of outside work per week negatively impacts GPA,^16,17^ so this value was used as the cutoff for time spent caring for family. PSS-10 items were analyzed individually using descriptive statistics. Reliability in our study using Cronbach’s α was very good for both faculty = 0.873 and students = 0.868.

## Results

Of 134 prospective student participants, 80 survey responses were received (response rate of 59.7%). One student skipped three items of the PSS10, and their total score was dropped; however, scores on the individual questions to which they responded have been included. For students, the average age of participants was 24 years (*SD* = 3.9). Gender distribution was 67.5% female and 28.8% male. The ethnic makeup of participants was 27.5% Asian American, 5.0% Black/African American, 13.8% Hispanic/Latino, 6.3% Middle Eastern, 2.5% mixed race, and 42.5% White/Caucasian. A minority of students (18.8%) were from out-of-state. Average GPA was reported as 3.5 (*SD* = 0.5). Participants take care of family members an average of 7.4 (*SD* = 13.8) hours per week, perform work outside of schoolwork 7.4 (*SD* = 9.4) hours per week, and sleep 6.3 (*SD* = 1.4) hours per day. The median total student loan debt was between $40,000 and $50,000.

Of the 53 faculty members invited to complete the survey, 44 responses were received (response rate of 83%). One faculty member skipped one item of the PSS10, and their total score was dropped; however, scores on nine individual questions to which they responded have been included. Two faculty members skipped answering all demographic and employment-related questions. Their total scores have been included for the overall mean PSS10 score for faculty, but not elsewhere in the analysis. Of the faculty respondents who answered demographic questions, the majority were female (79.49%), married (65.91%), teaching during the quarter in which the survey was administered (61.36%), tenured (50%), with a clinical practice site (54.54%), and had children <12 years (52.78%).

Faculty PSS10 scores ranged from 1 to 26, while student PSS10 scores ranged from 1 to 36, with higher scores indicating higher stress (**Figure 2A**). The mean PSS10 score for faculty was 15.50 (SD 5.84). The mean PSS10 score for students was 21.14 (SD 7.19). Thus, students had a higher perceived stress level than faculty, *t*(117) = -4.294, p<0.001. The mean PSS10 score for female faculty was 16.43 (SD 5.94) and 12.00 (SD 4.50) for male faculty. Analysis by gender indicated that female faculty’s perceived stress level was higher than that of male faculty *t*(36) = -2.301, p = 0.037. The mean PSS10 score for female students was 22.60 (SD 5.74) and 16.78 (SD 7.83) for male students. Analysis by gender indicated that female students’ perceived stress level is higher than that of male students, *t*(74) = -3.623, p = 0.001 (**Figure 2B**).

**Figure 2.**
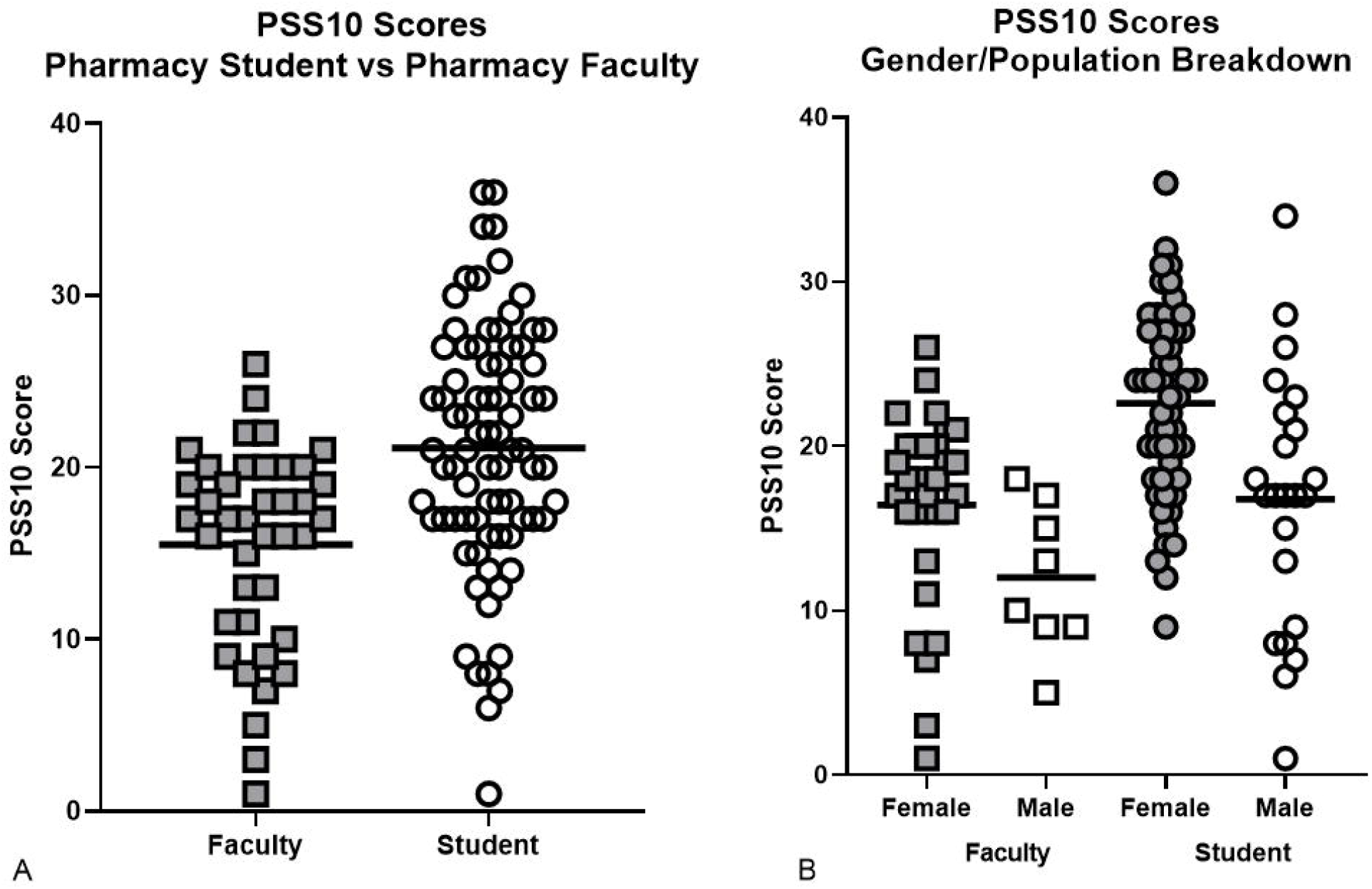
Average perceived stress PSS10 scores **A**. In pharmacy faculty (▄) versus first year pharmacy students (○) where samples include both males and females; and **B**. In female pharmacy faculty (▄) versus male pharmacy faculty (□) and in first year female pharmacy students (•) versus first year male pharmacy students (○). The lines (─) indicate the mean for each group.

The mean PSS10 score for female faculty with children <12 years of age was 18.85 (SD 4.41). Female faculty with no children were combined with those having children ≥12 years of age (due to small sample sizes), and the mean PSS10 score was 12.64 (SD 6.74). The mean PSS10 score for male faculty with children <12 years of age was 9.67 (SD 5.03). Male faculty with no children were combined with those having children ≥12 years of age (due to small sample sizes), and the mean PSS10 score was 13.40 (SD 4.04). Analysis by gender indicated that female faculty’s perceived stress level with children <12 years of age was higher than that of male faculty with children <12 years of age *t*(17) = -3.011, p = 0.008.

The mean PSS10 score for female students with ≥10 hours of family obligations was 22.71 (SD 5.25), while the mean of those with <10 hours of family obligations was 22.56 (SD 6.03). The mean PSS10 score for male students with ≥10 hours of family obligations was 12.80 (SD 8.87), while the mean of those with <10 hours of family obligations was 17.89 (SD 7.41). Analysis by gender indicated that female students’ perceived stress level with ≥10 hours of family obligations was higher than that of male students with ≥10 hours of family obligations *t*(18) = -3.223, p = 0.005.

We examined whether there were item differences between the various populations. When female faculty were compared to male faculty (**Figure 3**), significant gender differences were noted for Question 1 (p = 0.0422), Question 3 (p = 0.0467), Question 6 (p = 0.0103), and Question 10 (p = 0.0293). Similarly, when female students were compared to male students (**Figure 3**), significant gender differences were noted for Question 3 (p = 0.0068), Question 6 (p = 0.0389), Question 9 (p = 0.0224), and Question 10 (p = 0.0246).

**Figure 3.**
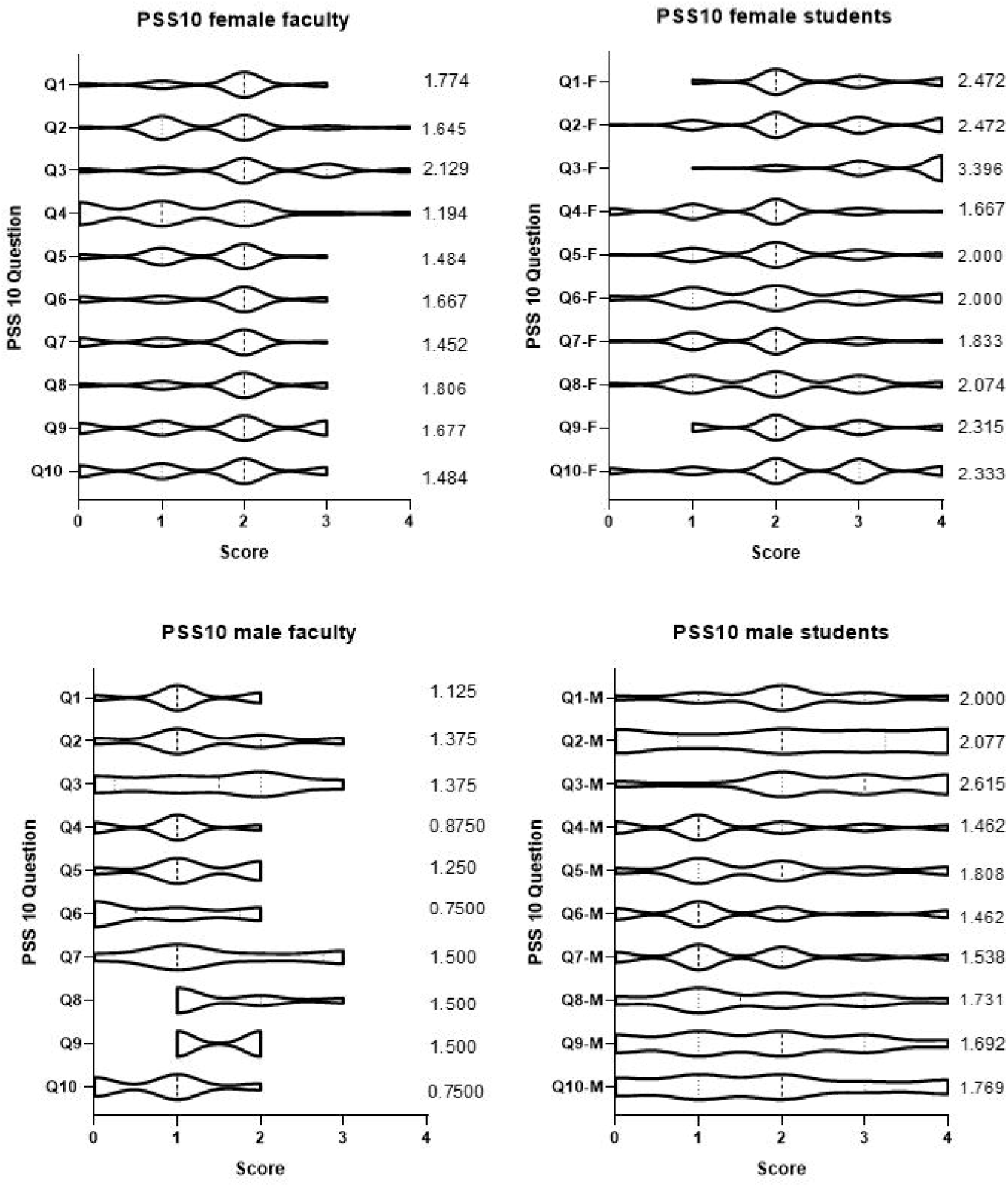
Violin plots depicting the distribution of PSS10 responses based on gender for pharmacy faculty and first year pharmacy students. The figures are scaled so that the width of each shape represents the density of scores at that value. The numbers provided to the right are the average scores for the individual question for that group.

When female faculty were compared to female students (**Figure 3**), significant differences were noted for seven items: Question 1 (p = 0.0055), Question 2 (p = 0.0006), Question 3 (p < 0.0001), Question 4 (p = 0.0465), Question 5 (p = 0.0303), Question 9 (p = 0.0117), and Question 10 (p = 0.0005). When male faculty were compared to male students (**Figure 3**), significant differences were noted for four items: Question 1 (p = 0.0269), Question 3 (p < 0.0064), and Question 10 (p = 0.0202).

There were no statistical differences in any PSS10 items for male versus female students if they reported <10 hours family time (**Figure 4**). However, we did find that some of the individual PSS10 questions are significantly different between male and female students if they reported ≥10 hours of family time (**Figure 4**). The individual questions that showed a significant difference between male and female students with ≥10 hours family time were Question 3 (p = 0.0225), Question 6 (p = 0.0423), Question 9 (p = 0.0201), and Question 10 (p = 0.0377). For female faculty with children <12 years of age versus those without children, or with those having children ≥12 years of age (**Figure 5**), the individual questions with statistical differences were Question 1 (p = 0.0092), Question 7 (p = 0.0069), and Question 8 (p = 0.0031). For male faculty with children <12 years of age versus male faculty without children, or with children ≥12 years (**Figure 5**), there was only one item where there are statistical differences, and that was Question 7 (p = 0.0438). When male faculty with children <12 are compared with female faculty with children <12 (**Figure 5**) there were significant differences for Question 1 (p = 0.0427), Question 3 (p = 0.0074), and Question 7 (p = 0.0195). For male versus female faculty without children or whose children ≥12 years of age (**Figure 5**), there were significant differences for only Question 7 (p = 0.0472).

**Figure 4.**
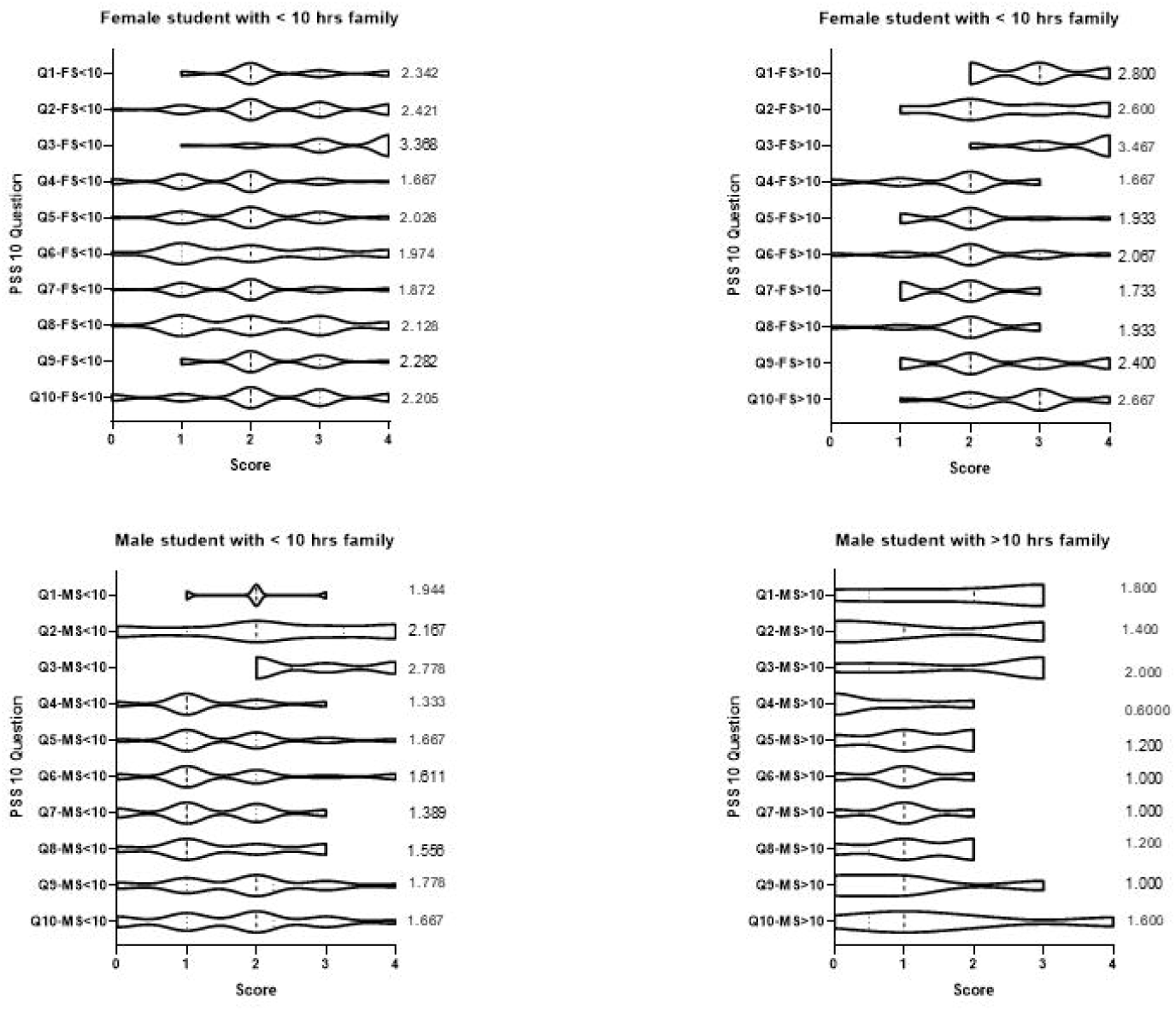
Violin plots depicting the distribution of PSS10 responses for first year pharmacy students based on gender and self-reported time spent on family responsibilities. The figures are scaled so that the width of each shape represents the density of scores at that value. The numbers provided to the right are the average scores for the individual question for that group.

**Figure 5.**
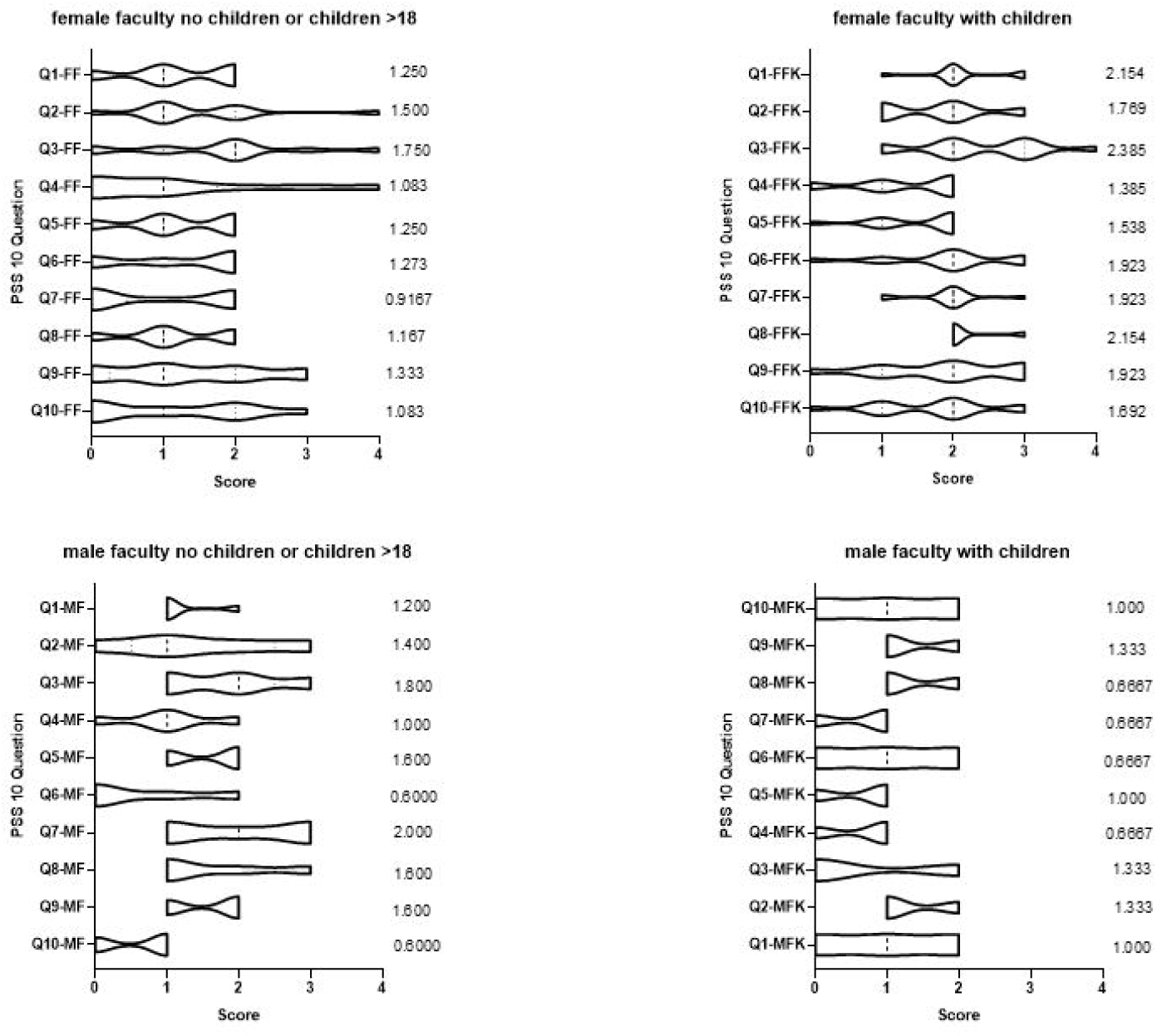
Violin plots depicting the distribution of PSS10 responses for pharmacy faculty based on gender and family responsibilities. The figures are scaled so that the width of each shape represents the density of scores at that value. The numbers provided to the right are the average scores for the individual question for that group.

## Discussion

Pharmacy students and faculty experience stress. Previous studies have not directly compared stress levels between pharmacy faculty and students. Others have shown no significant difference between undergraduate and graduate students’ stress levels.^18^ Similarly, older graduate entry medical students and their younger undergraduate counterparts have reported similar perceived stress profiles.^19^

Our survey results demonstrated pharmacy faculty have lower levels of perceived stress than pharmacy students. Several theoretical possibilities could contribute to a higher student perceived stress levels, including less developed coping behaviors, differences in available social support systems, and awareness of the root causes of stressors. This may be due to the likelihood that faculty have developed coping strategies over time. Additionally, a higher sense of control and fewer academic-related stressors might be present for faculty versus students.^1^

We examined whether personal characteristics, such as gender and family obligations, influence perceived stress levels between faculty and students. Similar to prior work, our study demonstrates female faculty and students report higher stress levels than male faculty and students. Many factors may be responsible for this difference. A recent study evaluated whether gender differences existed regarding career satisfaction, work-life balance, and pharmacy faculty stress nationally.^20^ Women faculty reported higher PSS10 scores than men (14.7 ± 6.6 vs. 11.8 ± 6.4, p < 0.001) even after controlling for factors such as age, marital status, number of children, academic rank, average hours worked per week, department, and type of institution. Women faculty in the aforementioned study reported less satisfaction with their current position and work-life balance than male faculty. A study by Lindfelt^21^ found that a greater proportion of women faculty reported a desire to leave their position versus male faculty (72.1% vs. 54.7%, p = 0.001). Furthermore, PSS10 scores for pharmacy faculty desiring to leave academia was significantly higher than the group planning to stay in academia (16.3 ± 6.7 vs. 13.0 ± 6.0, p < 0.001).

Similar to faculty, our study found female students reported higher levels of perceived stress than male students. An assessment of pharmacy students noted that female students had significantly higher PSS scores than male students.^22^ In an extensive nationwide PSS10 survey of first through fourth-year pharmacy students, females reported significantly higher stress levels than males.^3^ In a study by Beall, pharmacy students were surveyed during each year of the curriculum to identify perceived stress, stress triggers, and coping mechanisms.^23^ In this study, female first-year pharmacy students reported the highest degree of perceived stress, and female students reported higher stress with all listed triggers except one (completing surveys). In contrast, a recent study of first-year pharmacy students at three colleges of pharmacy reported a mean PSS10 score of 20.0 (SD 6.9) (n = 118), and gender was not predictive of perceived stress levels.^24^

Interestingly, our study found higher perceived stress for female faculty members with younger children but lower perceived stress for male faculty members. This may be due to the nature of parent-child interactions. Work by Musick^25^ demonstrated that mothers tend to spend more time as the primary caregiver performing structured, routine childcare tasks (e.g., cooking, cleaning, management activities, and solo parenting). In contrast, fathers tend to spend more time in a “helper” role in leisure activities with less responsibility, such as unstructured “play” time. In the Musick study, mothers also reported more sleep disturbances than fathers. Details of parental responsibilities were not collected in our research study but could be an interesting hypothesis to explore in the future. In line with these results, male students who spent >10 hours on family obligations had lower perceived stress than male students who spent <10 hours on family obligations. An explanation for this may lie in the nature of the family obligations as more demanding tasks may increase stress levels, whereas more social tasks may decrease stress levels. Also, of note, female college students reported an inability to concentrate in the classroom due to the demands at home and personal concern when they had to miss child activities due to class attendance.^26^ It is also important to note that while gender and child status accounted for 30.4% of the variance for faculty gender, family care accounted for only 15.1% of the variance for students suggesting other variables not accounted for in this study contribute to student stress. Based on our survey results and many others, colleges of pharmacy should invest resources to help reduce stress levels in both their faculty and student populations, and should develop focused strategies for females. Potential areas could include increased resources for supportive counseling, group support, targeted mentorship for females with similar lifestyles/demographics, and help with family/childcare. If such resources already exist, then colleges could analyze their level of utilization, the effect on stress reduction, and, if appropriate, the reasons for underutilization (lack of awareness, time, or anonymity) via focus groups or surveys.

Limitations of the study are attributed to the survey instrument and sampling methods. With a student response rate of 59.7%, there is a risk for non-response bias, with the population of students responding not reflecting the overall P1 student population. PSS10 results are subject to response bias as they rely on individuals self-reporting their perceived stress. Additionally, this study was performed at a private pharmacy school located in the Midwest, at a single point in time. Perceived stress and basic need satisfaction may vary throughout the academic year and over subsequent years of work and study for faculty and students, respectively. These measures might differ for publicly funded pharmacy schools or those located outside the Midwest region. Consequently, the results may not apply to other pharmacy schools. Only first-year professional students were surveyed in our study, and the results might have differed if P2, P3, and P4 students were included.

## Conclusion

We report perceived stress levels for first-year pharmacy students are higher than pharmacy faculty in our college. Perceived stress levels are higher with both female faculty and female students compared to their male counterparts. Family obligations impact female faculty and female students’ perceived stress levels to a greater extent than the male faculty and students. Targeted wellness interventions to alleviate stress surrounding family obligations should be considered on a college or university level. Additional studies on a larger level are needed to further elucidate distinct areas of stress for faculty and students that better inform us about which wellness interventions might be most successful.

## Data Availability

All data produced in the present work are contained in the manuscript

## Acknowledgments

The authors would like to acknowledge the faculty and students who participated in the surveys.

